# High Mortality Associated with Chikungunya Epidemic in Southeast Brazil, 2023

**DOI:** 10.1101/2024.07.03.24309912

**Authors:** Andre Ricardo Ribas Freitas, Antonio Silva Lima Neto, Erneson Alves Oliveira, José Soares Andrade, Luciano Pamplona Cavalcanti

## Abstract

The chikungunya virus (CHIKV) was first detected in Brazil in 2014 with two lineages, North Asia and ECSA (East/Central/South Africa). The ECSA lineage dominated, causing significant outbreaks, especially in densely populated areas. The co-circulation of dengue and chikungunya has complicated diagnosis and treatment. The 2023 chikungunya epidemic in Minas Gerais, Southeast Brazil, prompted an assessment of associated mortality using WHO methodologies.

The study used epidemiological data from the laboratory surveillance, disease notification and mortality systems of the Brazilian Ministry of Health. The North and Northeast Health Macroregions of Minas Gerais, with 2.5 million inhabitants, were analyzed. A Poisson regression model, excluding critical periods of COVID-19 and the chikungunya epidemic, estimated expected mortality. Excess deaths were calculated by comparing observed deaths with model estimates during the epidemic period.

During the epidemic, there were 890 excess deaths attributed to chikungunya, translating into a mortality rate of 35.1/100,000 inhabitants. The excess mortality rate was significantly 60 times higher than the deaths reported by surveillance, with only 15 deaths confirmed. The correlation between excess deaths and laboratory-confirmed chikungunya cases was strong, while the correlation with dengue and COVID-19 was negligible. The results highlighted the serious underestimation of chikungunya mortality by epidemiological surveillance.

We estimate that there were 890 excess deaths in the 2023 chikungunya epidemic in just one affected area of Minas Gerais, Brazil (2.5 million inhabitants). During the same year, only 420 chikungunya deaths were reported by all PAHO member countries. Epidemiological surveillance has underestimated the impact of this disease. Excess mortality is a crucial measure for understanding the impact of epidemics, as demonstrated by COVID-19 and influenza pandemics. The study highlights the need for complementary tools to traditional surveillance to better assess impacts on morbidity and mortality. Underreporting of chikungunya deaths has significant implications for public health responses and resource allocation. This study supports the importance of accurately quantifying mortality from emerging viruses to inform risk assessments and priority setting in public health interventions.

## Introduction

Until then free area, in Brazil in 2014 two lineages of the chikungunya virus (CHIKV) were detected, in the North, Asian lineage and in the Northeast, ECSA (East/Central/South Africa, CHIKV), with transmission sustained by Aedes aegypti. In the following years, only the ECSA lineage remained and caused several focal epidemics, mainly in the North and Northeast regions(1). In recent years there has been an increase in the number of cases with rapid territorial expansion, reaching densely populated areas in the South and Southeast regions. The co-circulation of dengue and chikungunya, associated with little clinical knowledge of the latter by health professionals, has made the diagnosis and treatment of the disease difficult, in addition to preventing a more reliable assessment of the impact of chikungunya in Brazil (2) and in other countries in the region.(3)

In the year 2023, Minas Gerais, a state located in the Southeast Region of Brazil, was the state most affected by the chikungunya epidemic(2). Using a methodology adopted by the World Health Organization(4), we assessed mortality associated with the chikungunya epidemic in the most affected areas

## Methods

We assessed the epidemiological situation of chikungunya based on cases available in the laboratory surveillance, disease notification and mortality systems maintained by the Brazilian Ministry of Health (http://tabnet.datasus.gov.br/). The North and Northeast Health Macro-regions of Minas Gerais were evaluated together because they are contiguous areas, similar from a social, epidemiological and climatic point of view(figure 1). Together, these Macro-regions total 2.5 million inhabitants, distributed between urban clusters, such as Montes Claros (largest city in the region, 413 thousand inhabitants) and extensive rural areas with low population density. The region’s climate is tropical and during the studied period there were no extreme weather events in the region, according to data from the National Institute of Meteorology (INMET, Appendix).

**Figure 1.**
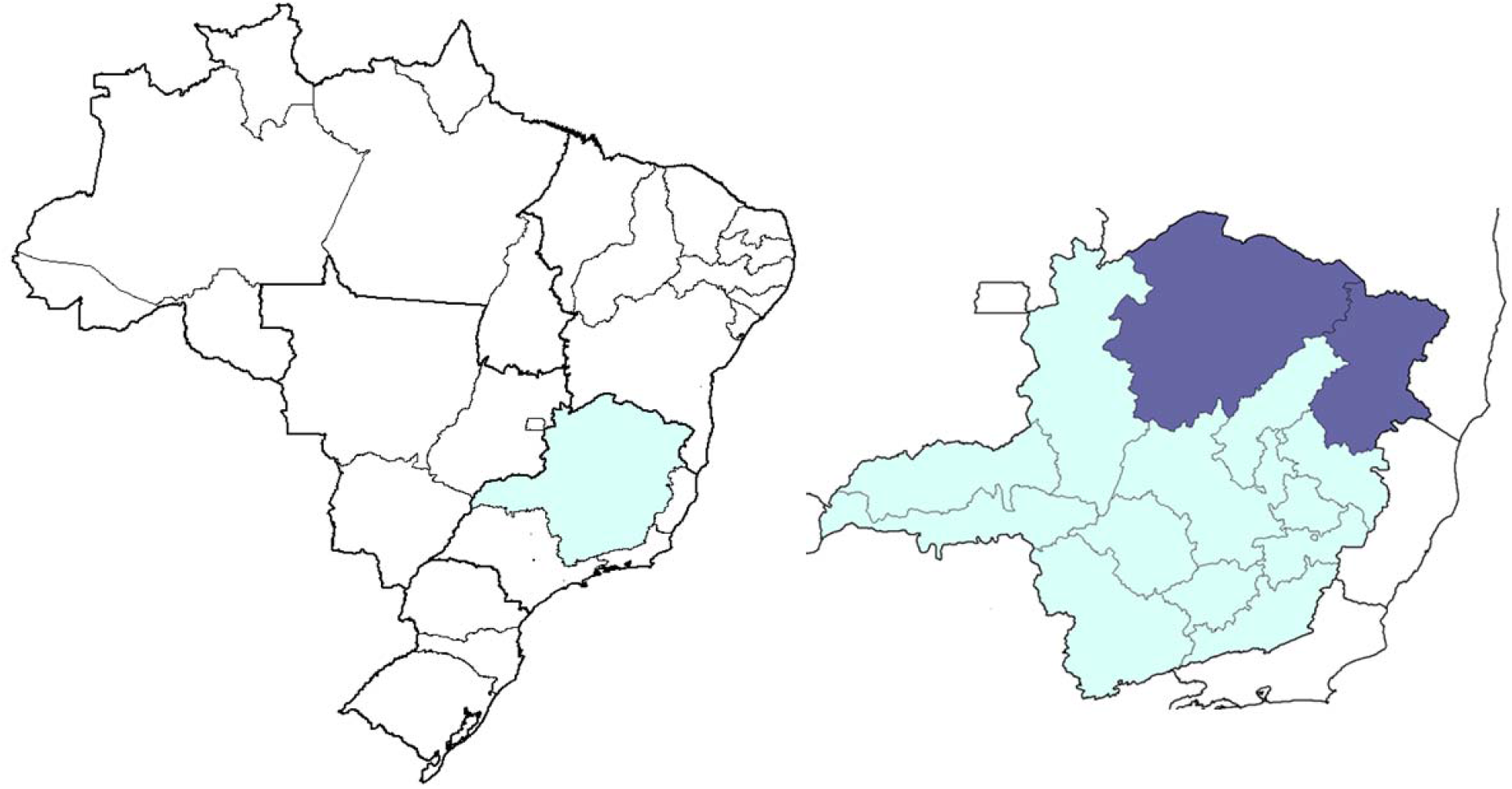
Map of Brazil highlighting the State of Minas Gerais and the North and Northeast Health Macro-regions

In table 1, we present the number of laboratory confirmed cases and calculate the coefficient of incidence and positivity of laboratory tests (positive tests/tests performed) of chikungunya, dengue and covid-19 (Severe Acute Respiratory Infection cases) for all Health Macro-regions of Minas Gerais, during the period of the epidemic. The regions selected for the present study had the highest positivity and incidences of chikungunya in the state and low positivity and incidences of dengue and covid-19.

**Table 1.**
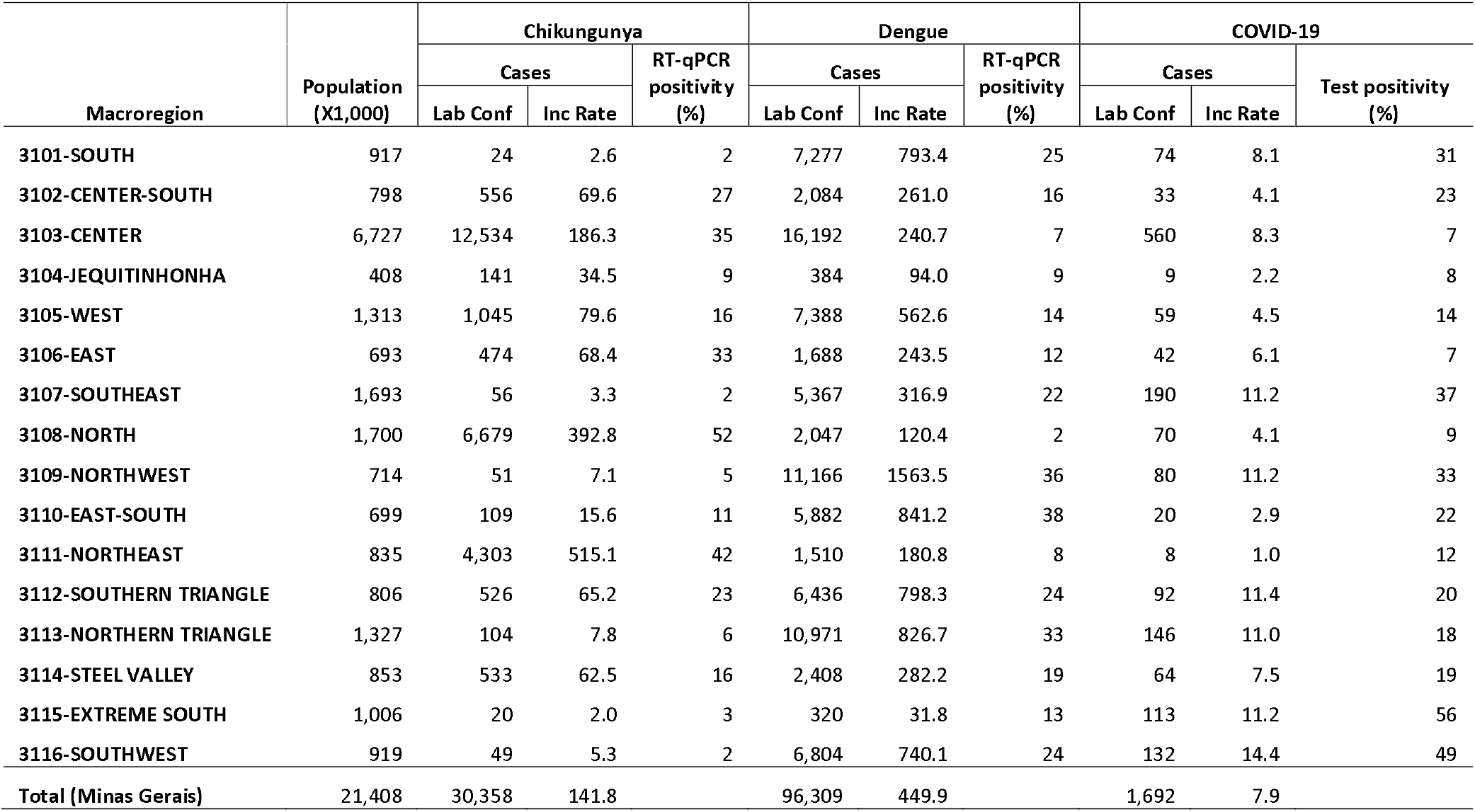
Population, laboratory confirmed cases, incidence rate and test positivity in the Health Macro-regions of Minas Gerais (chikungunya, dengue and covid-19, Dec 2022-May 2023, Brazil)

We assessed excess mortality associated with the 2023 chikungunya epidemic. A time series of average daily all-cause mortality was constructed for each month from January 2017 to December 2023. To estimate expected mortality for the period of chikungunya epidemic, a robust Poisson regression model was constructed based on the formula below:

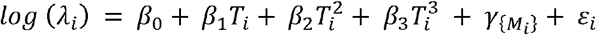

Where:

> λ_i_ represented the number of deaths in a particular month 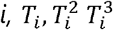 are time in months as a linear, quadratic and cubic to correct for secular fluctuations and 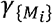 are the nominal variables associated with the months, to correct seasonal fluctuations and ϵ_i_ is the error term.

We estimated the regression coefficients excluding the most critical period of the covid-19 pandemic (Jan/2020-Jul/2022) and the period of the chikungunya epidemic (Dec/2022-May/2023). The period of the chikungunya epidemic was defined as the months with incidence coefficients above the 95% percentile of the analyzed period. Excess deaths were considered as the difference between observed deaths and those estimated by the model for the months in which observed deaths exceeded the upper limit of the 99% CI(4)(Figure 2).

**Figure 2.**
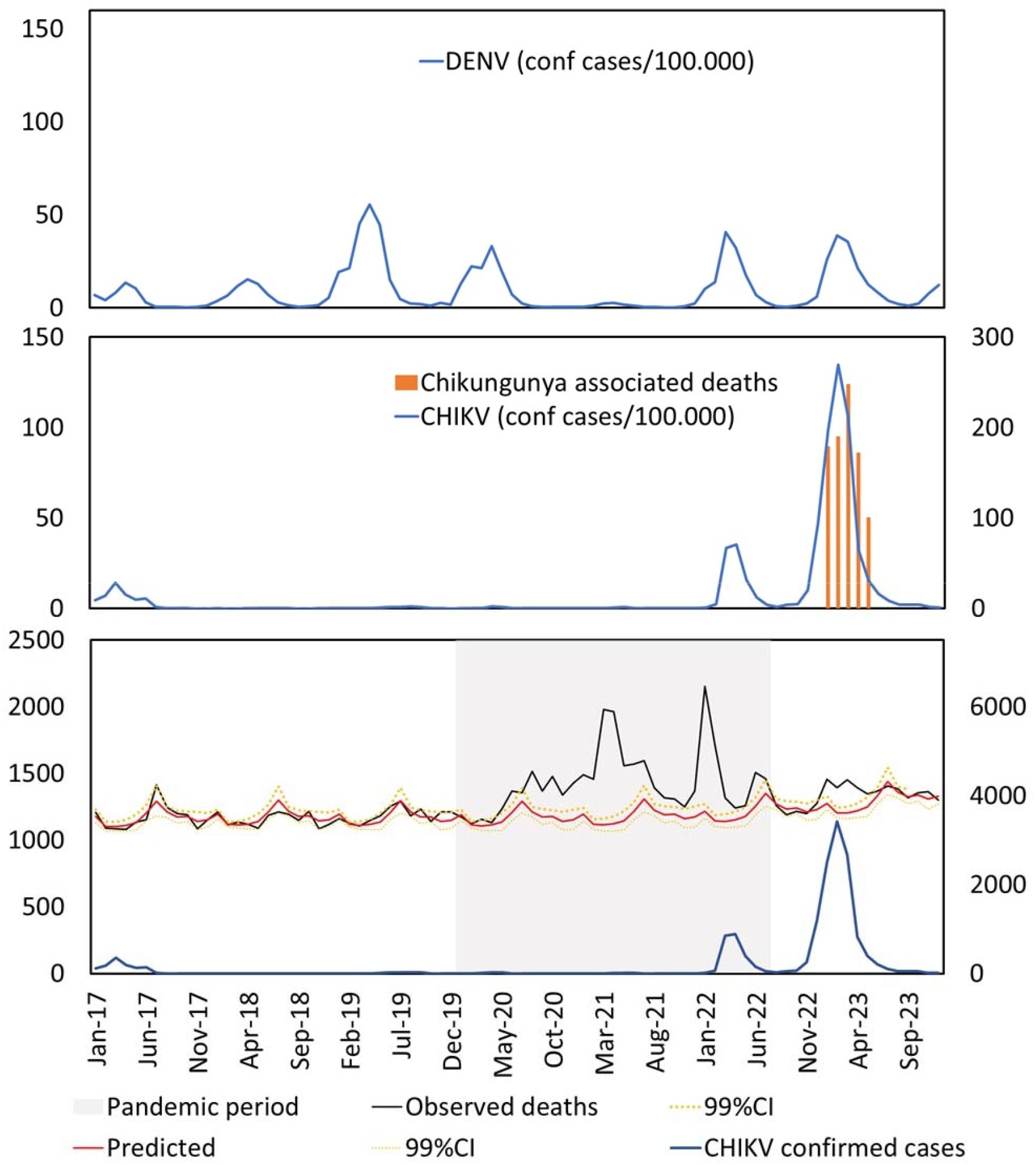
Bottom: absolute number of monthly cases of chikungunya (blue line, right scale), number of observed monthly deaths (black line, left scale), number of monthly deaths predicted by the model (solid red line, left scale) and upper and lower limits of the 99% confidence interval (dotted and solid yellow lines respectively, left scale). In the middle: monthly incidence coefficient of chikungunya per 100,000 inhabitants (blue line, left scale), monthly number of excess deaths (red columns, right scale). Above: dengue incidence coefficient per 100,000 inhabitants (blue line, left scale). All data refer to the North and Northeast Health Macro-regions of Minas Gerais.

## Results

During the chikungunya epidemic, there were 890 more deaths than the number expected for the period. This value is the mortality considered attributable to chikungunya. We also constructed a robust Poisson regression with sine and cosine seasonal components for Fourier decomposition, the results found were not statistically different (p=0.8) and we opted for the model presented due to its logical simplicity and smaller number of terms (Appendix)(5).

We found that the excess of monthly deaths has a significative Kendall’s tau-b correlation between the total number of confirmed monthly deaths, with laboratory-confirmed cases and with positivity by RT-qPCR test for chikungunya(table 2). On the other hand, there was a negative and non-significant correlation between the excess of monthly deaths and the total number of confirmed monthly deaths, confirmed cases and positive tests for covid-19.

**Table 2.**
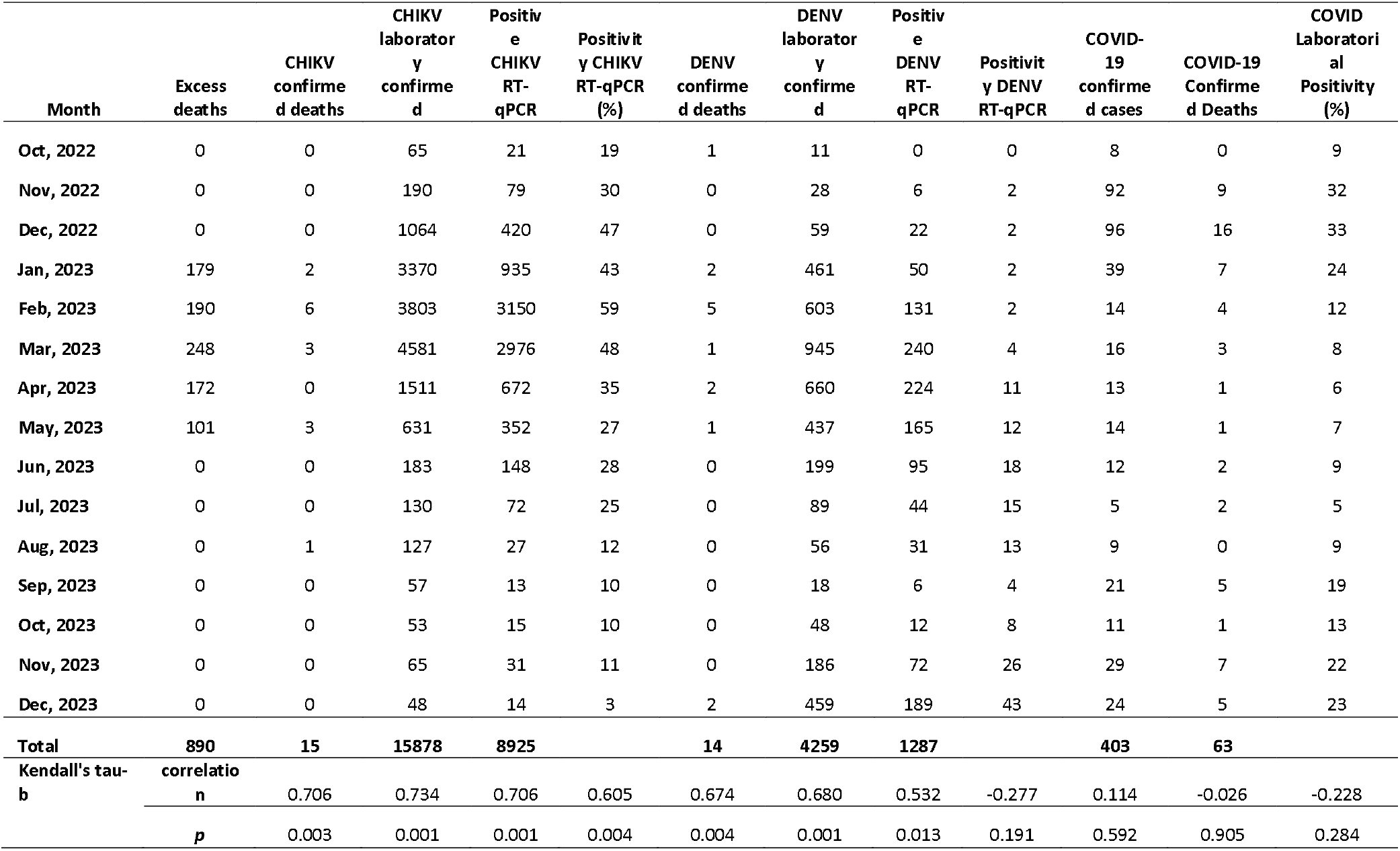
Excess deaths, laboratory-confirmed cases and deaths, incidence rate and test positivity per month North and Northeast Health Macro-regions of Minas Gerais (chikungunya, dengue and covid-19, Oct, 2022 - Dec, 2023, Brazil)

The correlation between excess monthly mortality and the number of dengue cases and deaths was an expected phenomenon due to the seasonal coincidence with chikungunya. However, there was a negative and non-significant correlation between excess monthly mortality and the positivity of laboratory tests (RT-qPCR) for dengue.

The excess mortality rate in the period was calculated at 35.1/100,000 inhabitants. This value is a little more than half the global average excess mortality rate from Covid-19 estimated by WHO for the first year of the pandemic (60/100,000) (4), it is seven times greater than the excess rate estimated global average mortality rate for the 2009 influenza pandemic (5/100,000 inhabitants) and is close to the excess mortality rate for the 1967 influenza pandemic (40/100,000)(6). In a similar study we found that in the 2016 chikungunya epidemic in Pernambuco there was an excess death rate of 47.9 and in the 2014 epidemic on the islands of Guadalupe and Martinique the excess deaths were 81.6 per 100,000 inhabitants(7,8). Mavalankar and colleagues calculated the excess mortality rate in Ahmedabab(India) during the 2006 chikungunya epidemic at 77.5 deaths per 100,000(3,800 deaths)(9) while no deaths have been officially confirmed in the country.

The number of deaths(890 deaths) attributable to the chikungunya epidemic is almost 60 times greater than the number of deaths(15 deaths) identified by epidemiological surveillance in the two Macro-regions studied. Despite representing a small fraction of the Brazilian territory, this excess of deaths observed is almost 8 times greater than the total number of deaths from chikungunya identified by epidemiological surveillance throughout Brazil in the year 2023(106 deaths). In Pernambuco(2016, Brazil) there were 4,505 excess deaths during the chikungunya epidemic period, but surveillance confirmed only 94 deaths. In Puerto Rico in the 2014 epidemic, only 31 deaths from chikungunya were confirmed, but there were 1,310 deaths higher than expected for the period(7,10). In these examples, the underestimation of the number of deaths by the official surveillance system was 98%, the same value found in the present study.

## Discussion

Covid-19 pandemic brought great interest to the topic of monitoring excess mortality as a fundamental tool for assessing the impact of emerging viruses, as it is applicable in different contexts and is less dependent on laboratory resources(4,6). Given the significant increase in the number of chikungunya cases in endemic areas and the expansion of this and other arboviruses to free areas, tools complementary to traditional epidemiological surveillance need to be added to better assess the impact in terms of morbidity and mortality.

Excess mortality is a measure that contributes to building solid evidence of the impact of epidemics. Influenza is a classic example of the use of this tool to more adequately measure the number of deaths, as a significant part of deaths occur due to bacterial complications, decompensation of pre-existing diseases, among others.(6)

Adequately quantifying the number of deaths caused by emerging viruses is a fundamental need so that risk assessment and priority-setting systems can direct resources to address epidemiological challenges.

In recent years, necropsy studies have demonstrated viral antigens and genetic material in tissues of important organs such as the CNS, heart, lung and liver(11–14). The fact that the worsening of chikungunya is caused by a severe systemic infection, immunological and hemodynamic dysfunction, associated with the dysfunction of multiple distinct organs makes it difficult to define a single clinical syndrome that characterizes the worsening, which makes clinical suspicion very difficult, especially as it is a disease emerging(11–14).

The first warning sign about the importance of severe forms of chikungunya was given in 2006 on the island of Reunion(15), the issue of underreporting of deaths was subsequently demonstrated in India (9). Since then, evidence of these warnings has been confirmed through different study designs, in different regions of the world, however, several researchers and even official documents still classify chikungunya disease as having a low risk of death.

This study did not receive funding and the authors declare that they have no conflicts of interest.

## Data Availability

All data produced in the present study are available upon reasonable request to the authors.

