## Supplement for "High Mortality Associated with Chikungunya Epidemic in Southeast Brazil, 2023"

**Data from meteorological station A506 Montes Claros-MG, obtained through the National Institute of Meteorology (INMET) website.** https://tempo.inmet.gov.br/GraficosAnuais/A001

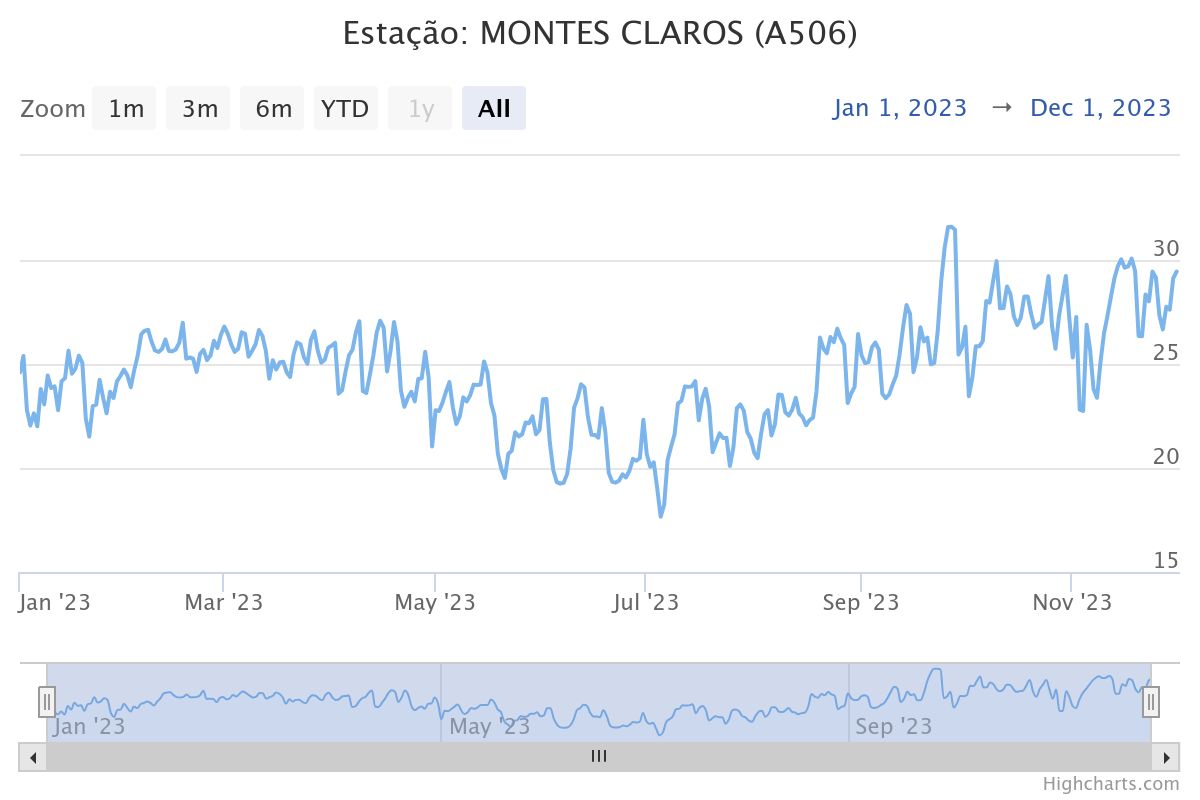

Figure 1 - Graph with temperature data from meteorological station A506 Montes Claros-MG, obtained through the National Institute of Meteorology (INMET) website. <https://tempo.inmet.gov.br/GraficosAnuais/A001>

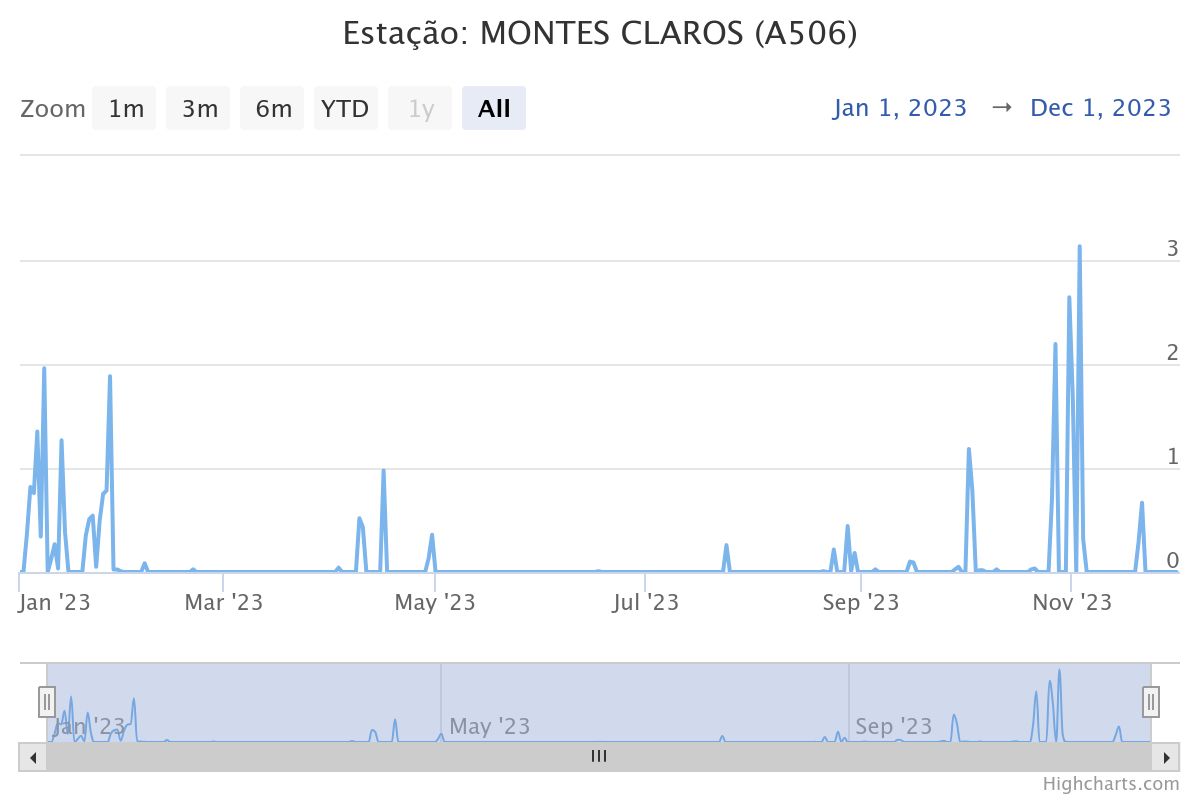

Figure 2 - Graph with precipitation data from meteorological station A506 Montes Claros-MG, obtained through the National Institute of Meteorology (INMET) website. <https://tempo.inmet.gov.br/GraficosAnuais/A001>

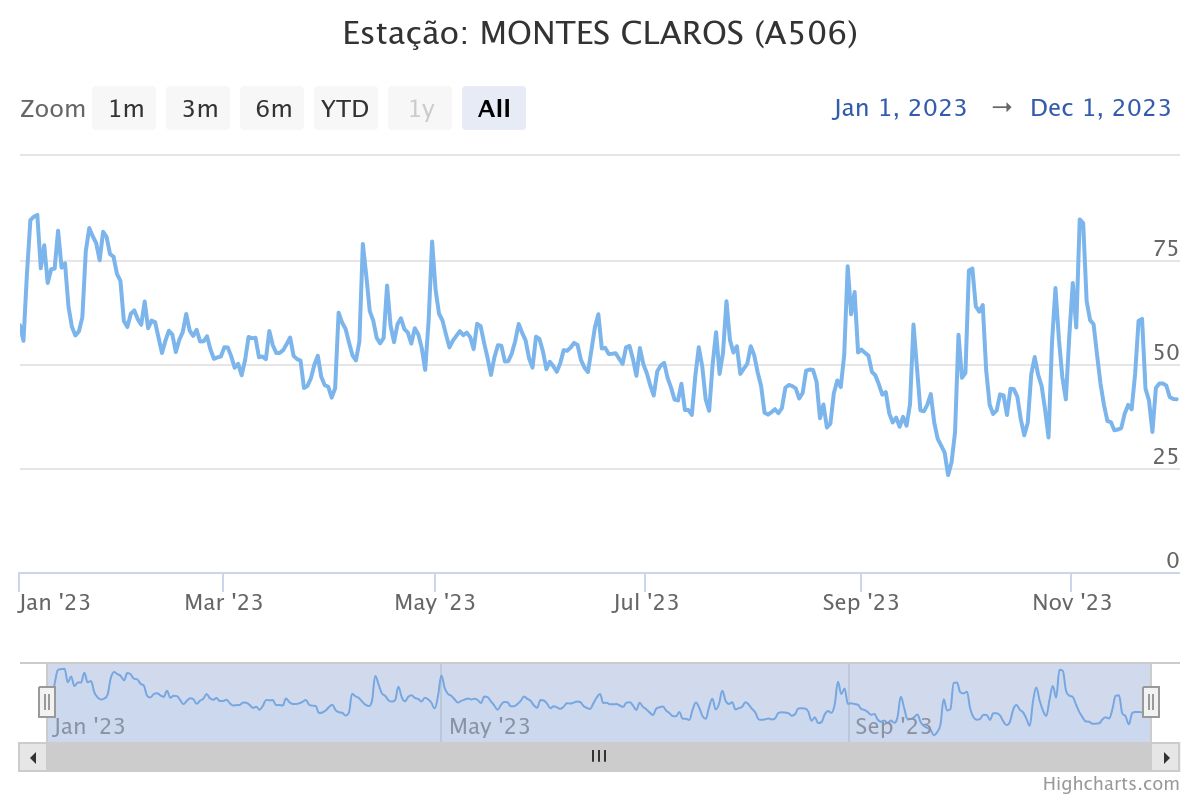

Figure 3 - Graph with humidity data from meteorological station A506 Montes Claros-MG, obtained through the National Institute of Meteorology (INMET) website. <https://tempo.inmet.gov.br/GraficosAnuais/A001>

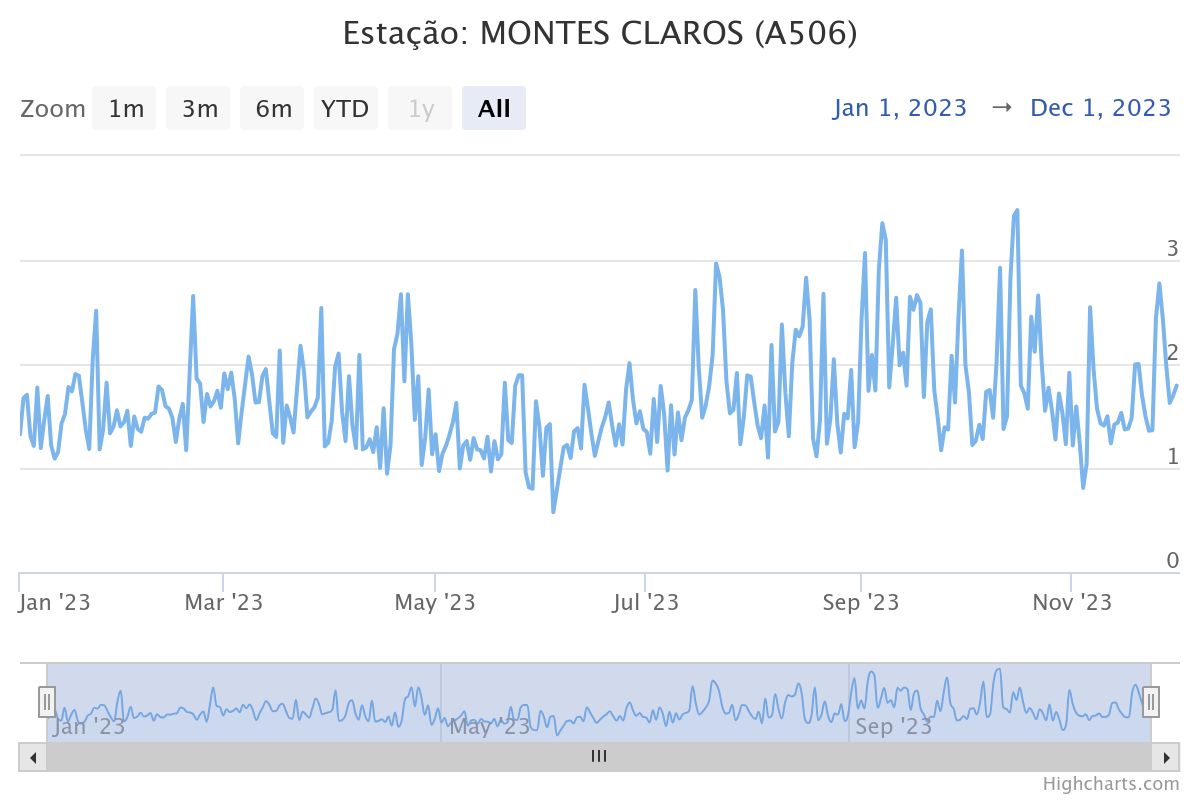

Figure 4 - Graph with wind velocity data from meteorological station A506 Montes Claros-MG, obtained through the National Institute of Meteorology (INMET) website. <https://tempo.inmet.gov.br/GraficosAnuais/A001>

**OUTPUT OF FINAL MODEL**

| **Quality of fit^a^** | | | |
| --- | --- | --- | --- |
|  | Valor | gl | Valor/gl |
| Deviance | 61,990 | 34 | 1,823 |
| Pearson qui-square | 62,294 | 34 | 1,832 |
| Log-likelihood | -249,660 |  |  |
| AIC | 529,320 |  |  |
| AICC | 543,866 |  |  |
| Critério de informações Bayesiano (BIC) | 557,698 |  |  |
| AIC consistente (CAIC) | 572,698 |  |  |
| Model: (Intercept), Month, LINEAR, SQUARE, CUBE | | | |

| Parameter | B | Error | 99% Wald CI | | Test | | |
| --- | --- | --- | --- | --- | --- | --- | --- |
|  |  |  | Lower | Upper | Qui-square Wald | gl | Sig. |
| (Intercept) | 7,056 | ,0205 | 7,003 | 7,109 | 118332,940 | 1 | ,000 |
| [Mon=Apr ] | -,049 | ,0191 | -,098 | ,000 | 6,572 | 1 | ,010 |
| [Mon=Ago ] | ,032 | ,0167 | -,011 | ,075 | 3,674 | 1 | ,055 |
| [Mon=Dec ] | -,020 | ,0223 | -,078 | ,037 | ,831 | 1 | ,362 |
| [Mon=Feb ] | -,051 | ,0157 | -,092 | -,011 | 10,609 | 1 | ,001 |
| [Mon=Jan ] | ,014 | ,0185 | -,034 | ,062 | ,581 | 1 | ,446 |
| [Mon=Jul ] | ,098 | ,0313 | ,017 | ,179 | 9,803 | 1 | ,002 |
| [Mon=Jun ] | ,028 | ,0212 | -,026 | ,083 | 1,759 | 1 | ,185 |
| [Mon=Mai ] | -,032 | ,0245 | -,095 | ,031 | 1,697 | 1 | ,193 |
| [Mon=Mar ] | -,056 | ,0171 | -,100 | -,012 | 10,720 | 1 | ,001 |
| [Mon=Nov ] | -,028 | ,0248 | -,092 | ,035 | 1,316 | 1 | ,251 |
| [Mon=Oct ] | ,003 | ,0171 | -,041 | ,047 | ,025 | 1 | ,874 |
| [Mon=Sep ] | 0^a^ | . | . | . | . | . | . |
| LINEAR | ,002 | ,0016 | -,002 | ,006 | 1,677 | 1 | ,195 |
| SQUARE | -8,431E-5 | 4,9898E-5 | ,000 | 4,422E-5 | 2,855 | 1 | ,091 |
| CUBE | 9,968E-7 | 4,2800E-7 | -1,057E-7 | 2,099E-6 | 5,424 | 1 | ,020 |
| (Escala) | 1^b^ |  |  |  |  |  |  |
| Model: (Intercepto), Mes, LINEAR, SQUARE, CUBE | | | | | | | |

**OUTPUT OF ALTERNATIVE MODEL**

Model using sine and cosine terms in the model to control for seasonality, regression^1^ formula can written as:

log(y)=β0+β1t+β2t^2^+β3sin(2πt12)+β4cos(2πt12)+β5sin(4πt12)+β6cos(4πt12)+β7sin(8πt12)+β8cos(8πt12)

Where:

- β0 is the intercept.
- β1 is the coefficient for the time in months.
- β2 is the coefficient for the time squared.
- β3 and β4 are the coefficients for the annual seasonal components.
- β5 and β6 are the coefficients for the doubled frequency seasonal components.
- β7 and β8 are the coefficients for the quadrupled frequency seasonal components.

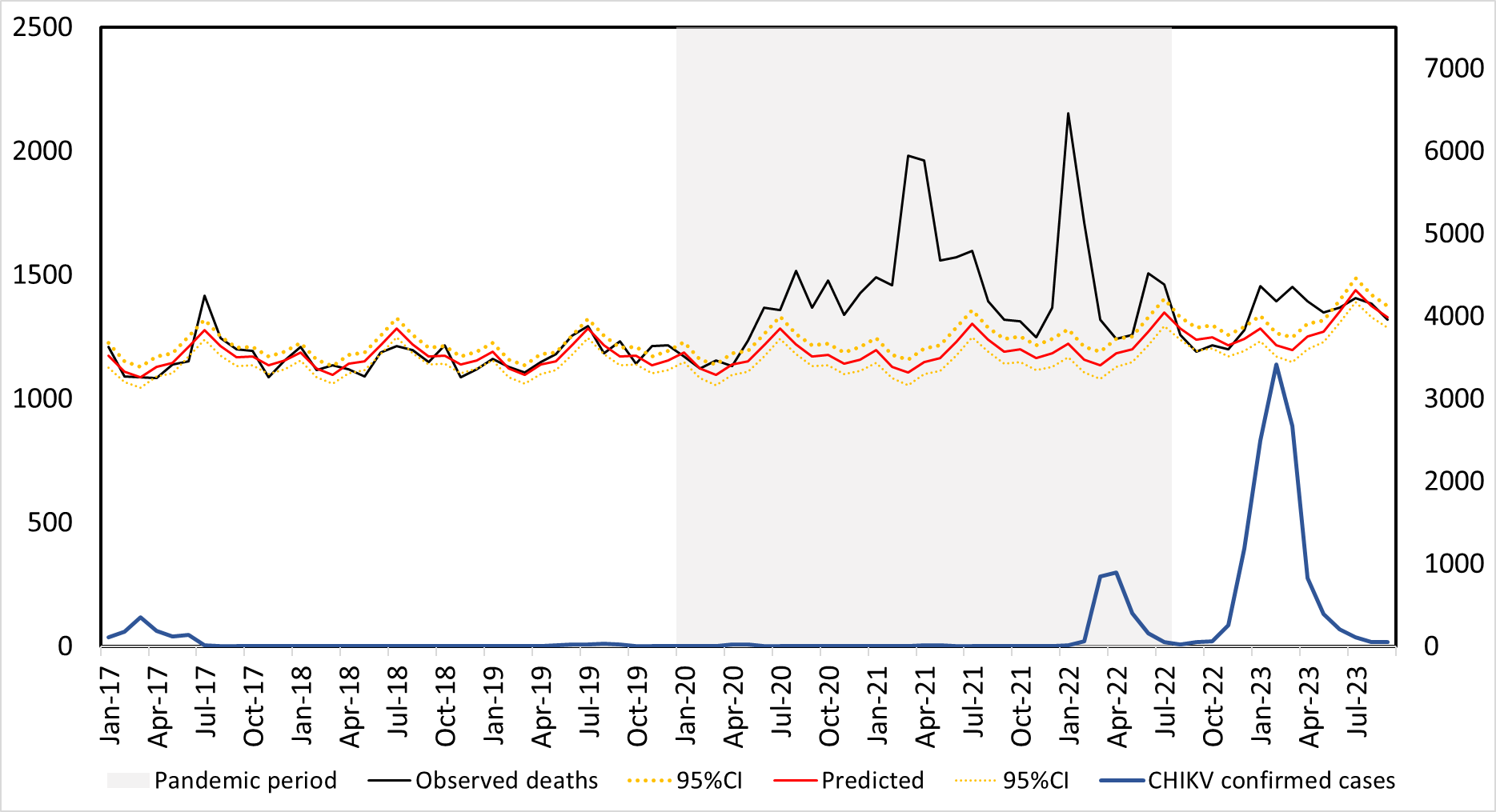

| **Quality of fit^a^** | | | |
| --- | --- | --- | --- |
|  | Valor | gl | Valor/gl |
| Deviance | 64,089 | 39 | 1,643 |
| Qui-quadrado de Pearson | 64,552 | 39 | 1,655 |
| Log da Verossimilhança^b^ | -250,709 |  |  |
| AIC | 521,419 |  |  |
| AICC | 527,208 |  |  |
| BIC | 540,337 |  |  |
| CAIC | 550,337 |  |  |
| Model: (Intercept), LINEAR, SQUARE, CUBE, SIN, COS, SIN2, COS2, SIN4, COS4 | | | |

| Parameter | B | Error | 99% Wald CI | | Test | | |
| --- | --- | --- | --- | --- | --- | --- | --- |
|  |  |  | Lower | Upper | Qui-square Wald | gl | Sig. |
| (Intercept) | 7,051 | ,0155 | 7,012 | 7,091 | 207090,594 | 1 | ,000 |
| LINEAR | ,002 | ,0016 | -,002 | ,006 | 1,647 | 1 | ,199 |
| SQUARED | -8,389E-5 | 4,9807E-5 | ,000 | 4,441E-5 | 2,837 | 1 | ,092 |
| CUBE | 9,963E-7 | 4,3113E-7 | -1,143E-7 | 2,107E-6 | 5,340 | 1 | ,021 |
| SIN | -,020 | ,0068 | -,038 | -,003 | 9,057 | 1 | ,003 |
| COS | -,037 | ,0074 | -,057 | -,018 | 25,737 | 1 | ,000 |
| SIN2 | -,004 | ,0064 | -,021 | ,012 | ,454 | 1 | ,500 |
| COS2 | ,035 | ,0070 | ,017 | ,053 | 24,922 | 1 | ,000 |
| SIN4 | -,004 | ,0066 | -,021 | ,013 | ,322 | 1 | ,570 |
| COS4 | ,023 | ,0070 | ,005 | ,041 | 10,574 | 1 | ,001 |
| (Escale) | 1^a^ |  |  |  |  |  |  |
| Model: (Intercept), LINEAR, SQUARED, CUBE, SIN, COS, SIN2, COS2, SIN4, COS4 | | | | | | | |

Qui-square of Likelihood Ratio Test: p = 0.8, p-value is greater than 0.05 indicating that there is no statistically significant difference between the models. We prefer model 1 due to its logical simplicity and smaller number of parameters.

1-European Centre for Disease Prevention and Control. Trend analysis guidance for surveillance data [Internet]. Stockholm; 2024. Available from: https://www.ecdc.europa.eu/sites/default/files/documents/trends-analysis-guidance-jan23.pdf
